# A 10-year retrospective analysis of *Toxoplasma gondii* qPCR screening in allogeneic hematopoietic stem cell transplantation recipients

**DOI:** 10.1101/2022.06.24.22276811

**Authors:** Alienor Xhaard, Alban Villate, Samia Hamane, David Michonneau, Jean Menotti, Marie Robin, Flore Sicre de Fontbrune, Nathalie Dhédin, Régis Peffault de la Tour, Gérard Socié, Stéphane Bretagne

**Affiliations:** Service d’hématologie-greffe, Hôpital Saint-Louis, APHP, Université Paris Diderot, Paris, France; Service d’hématologie et thérapie cellulaire, CHRU Tours, Tours, France; Laboratoire de mycologie-parasitologie, Hôpital Saint-Louis, APHP, Université Paris Diderot, Paris, France; INSERM UMR 976 (Team Insights), Université Paris Diderot, Paris, France; Laboratoire de mycologie-parasitologie, Hospices Civils de Lyon, Lyon, France; Service d’hématologie adolescents jeunes adultes, Hôpital Saint-Louis, APHP, Université Paris Diderot, Paris, France; Université Paris Cité

## Abstract

Weekly blood *Toxoplasma gondii* DNA screening using real-time quantitative polymerase chain reaction (qPCR) has been implemented in all allogeneic hematopoietic stem cell transplantation (alloHSCT) recipients at our hospital. We retrospectively analyzed the consequences of a positive blood qPCR in the management of *Toxoplasma* infection (TI) and disease (TD).

From 2011 to 2020, 52 (4.13%) of 1 257 alloHSCT recipients had at least one positive qPCR, 45 (3.5%) with TI and seven (0.56%) with TD (central nervous system involvement). Forty-four patients were qPCR-positive before day 100, 30 without and 14 with anti-*Toxoplasma* prophylaxis. Twenty-five of them (56.8%) started or continued prophylactic dosage treatment: all became qPCR-negative, including 20 (80%) receiving only prophylactic dosage treatment. Twenty-four of them (54.5%) received non-prophylactic dosage treatment: qPCR became negative in 22/24 (91.7%), while TI contributed to death in two cases. Six of the eight patients diagnosed after D100 were breakthrough TI or TD. No death was attributable to TI or TD. *qPCR* kinetics available for 24 patients increased until anti-*Toxoplasma* treatment began, then decreased with all treatment regimens.

Clinical follow-up and qPCR monitoring with quantification of the parasitic load appears a reasonable strategy to avoid TD and to use minimal effective dosage of anti-*Toxoplasma* treatments.

## Introduction

Toxoplasmosis is a zoonosis caused by *Toxoplasma gondii*, an intracellular coccidian parasite with numerous warm-blood intermediate hosts, and cats as the definite host. In humans, primary infection occurs after ingestion of cysts-containing undercooked meat or oocysts from the environment. Primary infection, usually asymptomatic, leads to the latency of bradyzoites in astrocytes, retina cells and muscles. In immunocompromised patients, toxoplasmosis mainly results from reactivation of latent cysts and its incidence depends on geographical endemicity, highly dependent on culinary habits.

Toxoplasmosis in allogeneic hematopoietic stem cell transplantation (alloHSCT) recipients remains a matter of concern (1). Prophylaxis using trimethoprim/sulfamethoxazole (TMP/SMX) is recommended in seropositive patients (2,3), but usually postponed until engraftment, for fear of possible hematological toxicity. Most cases occur between day 20 (D20) and D180 post-alloHSCT (4–6), but later or sooner occurrences have been reported (7). Outcomes were particularly dismal before the 2000s, with an estimated attributable mortality rate over 50%, probably due to a late diagnosis (8–10). Early *Toxoplasma* detection, using real-time quantitative polymerase chain reaction (qPCR) monitoring, has enabled the incidence of *Toxoplasma* disease and subsequent mortality to decrease. Indeed, circulating DNA detection usually precedes symptoms by 14-16 days (4,6,11). However, qPCR screening leads to the observation of qPCR-positive poorly symptomatic or asymptomatic patients: clinicians can then be reluctant to prescribe drugs possibly responsible for hematological toxicity (TMP/SMX or other sulfonamides) (12). Moreover, it is not yet known if all qPCR-positive patients will develop *Toxoplasma* disease in the absence of specific treatment, and there are no data and no recommendations on the best therapeutic option for asymptomatic patients with positive qPCR.

In our center, a strategy of weekly *Toxoplasma* qPCR screening of all alloHSCT recipients has been implemented for more than 10 years. The aims of this observational monocentric retrospective study were to describe the epidemiology of toxoplasmosis in alloHSCT recipients, and anti-*Toxoplasma* treatment in qPCR-positive patients.

## Patients and methods

### Patients and definitions

The study was conducted in the JACIE- (Joint Accreditation Committee for International Society for Cell and Gene Therapy (ISCT) Europe) and EBMT- (European Society for Blood and Marrow Transplantation) accredited hematology transplantation ward at Saint-Louis Hospital (Paris, France). AlloHSCT recipients transplanted between January 2011 and December 2020 were included. Data were collected from the PROMISE database and local medical records.

*Toxoplasma* infection (TI) and disease (TD) were defined according to the European Society for Blood and Marrow Transplant-Infectious Diseases Working Party guidelines (4,13). TI was defined as ≥ one positive PCR test from blood in a patient without evidence of organ involvement, with or without fever. TD was defined as definite (histological evidence), probable (clinical and radiological evidence plus ≥ one positive PCR test from blood, cerebrospinal fluid (CSF) or bronchoalveolar lavage fluid (BAL), and absence of other pathogen), or possible (suggestive imaging and response to specific treatment, but no laboratory evidence). The day of the first positive qPCR was defined as the day of TI or TD diagnosis. Mortality was considered toxoplasmosis-attributable if it occurred with positive qPCR or compatible clinical signs, and in the absence of alternative diagnosis. Toxoplasmosis was considered as a death-contributing factor if qPCR remained positive while the patient died for another identified cause (13).

### Microbiological and diagnostic investigations

Prior to alloHSCT, an anti-*Toxoplasma* immunoglobulin-G (IgG) assay was systematically performed (Platelia Toxo IgG TMB BIORAD until 2014 and ARCHITECT Toxo IgG thereafter) in donors and recipients (recipient within three months and donor within 28 days, respectively). The in-house qPCR targeted the *Toxoplasma*-specific AF 146527 DNA sequence (14) with an internal control (Simplexa™ Extraction & Amplification Control, Focus Diagnostics). qPCR monitoring was performed weekly on venous blood samples (1 ml), regardless of serological status from alloHSCT until D100, and thereafter at the discretion of the physician in cases of graft-versus-host-disease (GVHD), prolonged immunosuppression, or hospitalization (2,15). qPCR results were expressed as a quantification cycle (Cq) (16), with a low parasitic load corresponding to Cqs between 36 and 45. The laboratory subscribed to an independent external quality control for both *Toxoplasma* serology (Centre Toulousain pour le contrôle de qualité en Biologie clinique, France) and qPCR (Quality Control for Molecular Diagnostics, Glasgow, Scotland, United Kingdom).

In cases of positive *Toxoplasma* qPCR, the decision over whether to perform a diagnostic work-up was made by the physician in charge and usually included a central nervous system (CNS) computed-tomography (CT-scan) or magnetic-resonance imaging, thorax and abdomen CT-scans and fundus ophthalmoscopy. Blood control qPCR was performed once to twice weekly after the first positive sample.

### Anti-Toxoplasma prophylaxes and treatment (**Table 1**)

*Toxoplasma gondii* and *Pneumocystis jirovecii* prophylaxis was given to all patients after engraftment. Prophylaxis in our center was pyrimethamine/sulfadoxine (P/So) until commercialization ceased in 2017 (15,17). From then on, the most used prophylaxis has been TMP/SMX. However, atovaquone was preferentially used in patients with cytopenia, due to concerns about the possible hematological toxicity of TMP/SMX (12). The therapeutic option was led by the treating physician. Since qPCR was performed as a screening test before D100 and according to clinical suspicion later, we separated the two situations for further analyses.

**Table 1.** Molecules and dosages used for *Toxoplasma* prophylaxis and treatment.

| Dosage / treatment | Prophylactic | Non-prophylactic |  |
| --- | --- | --- | --- |
|  |  | Intermediate | High |
| TMP/SMX | 400 mg/d or 800 mg/d given three days/week | 800-1 600 mg/d | 2 400-4 800 mg/d |
| P/So | 1 500 mg/75 mg once weekly |  |  |
| Atovaquone | 750 mg bid |  |  |
| P/Sa |  |  | P: 50-100 mg<br>Sa: 4 000-6 000 mg/d |

Abbreviations: d: day; mg: milligrams; P: pyrimethamine; Sa: sulfadiazine; So: sulfadoxine;
TMP/SMX: trimethoprim/sulfamethoxazole

### Ethical consideration

The Institutional Review Board approved this study of the Saint-Louis Hospital. All patients gave their written consent for data collection. The study followed the Declaration of Helsinki.

### Statistical analyses

Continuous variables are expressed as median [IQR] and compared using Dunn’s test with Bonferroni correction. Categorical variables are expressed as frequencies (percentages) and compared using Fisher’s test. All analyses were performed using SAS version 9.4.6 (SAS Institute Inc., Cary, NC, USA). A two-tailed significance level p<0.05 was used.

## Results

### Study population and toxoplasmosis incidence

During the 10-year study period, 1 258 alloHSCTs were performed, including 1 257 donor/recipient (D/R) couples with available pre-alloHSCT *Toxoplasma gondii* serology. Anti-*Toxoplasma* IgG were positive in 734 recipients (58.3%) and 379 donors (30.1%). Excluding 65 cord blood alloHSCTs, anti-*Toxoplasma* IgG were positive in 264/579 (45.6%) of related and in 114/613 (18.6%) of unrelated adult donors. The D/R combinations were 272 (21.6%) D+/R+, 462 (36.8%) D-/R+, 107 (8.5%) D+/R-, and 416 (33.1%) D-/R-. The median age of the 1 257 patients was 45.6 years [IQR: 28-58]: 31 [IQR: 21-46] for *T. gondii* seronegative patients and 53 [IQR: 40-55] for seropositive patients. Overall mortality at D100 and one year after alloHSCT were 135/1 257 (10.7%) and 309/1 257 (24.6%) for the entire cohort, and 127/1 205 (10.5%) and 291/1 205 (24.1%) for the qPCR-negative patients, respectively.During the study period, 22 154 qPCR tests were performed (median/patient: 15 [IQR: 9-21]) without significant variation according to the year of sampling. In total, 52 patients (4.13%) had at least one *T. gondii* positive qPCR, i.e. a mean of five cases a year.

### *Characteristics of the 52* T. gondii *positive qPCR patients (Table 2 and supplementary Table 1*)

Thirty-one patients were male (56.9%), and median age at alloHSCT was 42 [IQR: 16-52]. Before alloHSCT, five patients were anti-*Toxoplasma* IgG negative. Two seropositive patients had a previous *Toxoplasma* reactivation before alloHSCT (one qPCR-positive during a septic shock treated by TMP/SMX and one ocular toxoplasmosis treated by atovaquone, seven and three months before alloHSCT, respectively). Forty-five patients had TI and seven had CNS TD (3.5% and 0.56% of 1 257 alloHSCT recipients, respectively). The seven TD were probable (CSF qPCR was positive in the two lumbar punctures performed) and the positive blood qPCR anticipated the diagnosis of cerebral involvement in all cases. Twenty-two (42.3%) patients died a median of 155 days (IQR [77-253]) after alloHSCT: toxoplasmosis contributed to death in two TI and one TD, respectively. In patients diagnosed before D100, the overall mortality rate was 43% (19/44), and 64% (9/14) in patients with anti-*Toxoplasma* prophylaxis.

**Table 2:** Characteristics of the 52 *T. gondii* positive qPCR patients according to the time of qPCR-positivity and the presence or not of prophylaxis at the first qPCR-positive result.

|  | Total | ≤ D100, w/o prophylaxis | ≤ D100 with prophylaxis | > D100 |
| --- | --- | --- | --- | --- |
| **Number of episodes** | 52 | 30 (57.7) | 14 (26.9) | 8 (15.4) |
| **Patients features, n (%)** |  |  |  |  |
| Male gender, n (%) | 31 (59.6) | 16 (53.3) | 10 (71.4) | 5 (62.5) |
| Median age at alloHSCT, yrs [IQR] | 42 [28-52] | 42 [25-52] | 40 [30-52] | 46 [36-51] |
| Underlying hematological condition |  |  |  |  |
| Acute myeloid leukemia | 21 (40.4) | 10 (33.3) | 6 (42.9) | 5 (62.5) |
| Acute lymphoblastic leukemia | 8 (15.4) | 5 (16.7) | 2 (14.3) | 1 (12.5) |
| Myelodysplastic syndrome | 4 (7.7) | 3 (10) | 1 (7.1) | 0 |
| Myeloproliferative neoplasm | 9 (17.3) | 8 (26.7) | 1 (7.1) | 0 |
| Non-Hodgkin lymphoma | 2 (3.8) | 0 | 1 (7.1) | 1 (12.5) |
| Hodgkin lymphoma | 1 (1.9) | 1 (3.3) | 0 | 0 |
| Sickle cell disease | 3 (5.8) | 2 (6.7) | 1 (7.1) | 0 |
| Others* | 4 (7.7) | 1 (3.3) | 2 (14.3) | 1 (12.5) |
| **D / R *T. gondii* IgG, n (%)** |  |  |  |  |
| D unknown / R negative | 1 (1.9) | 0 | 0 | 1 (12.5) |
| D positive / R positive | 7 (13.5) | 5 (16.7) | 1 (7.1) | 1 (12.5) |
| D negative / R positive | 40 (76.9) | 25 (83.3) | 10 (71.4) | 5 (62.5) |
| D positive / R negative | 2 (3.8) | 0 | 1 (7.1) | 1 (12.5) |
| D negative / R negative | 2 (3.8) | 0 | 2 (14.3) | 0 |
| **Positive *T. gondii* qPCR results** |  |  |  |  |
| Median number, n [IQR] | 2 [1-4] | 3 [2-4] | 1.5 [1-4] | 1.5 [1-2] |
| Median days from alloHSCT to first positive qPCR, [IQR] | 26 [17-60] | 19 [14-25] | 48 [28-61] | 202 [163-282] |
| D0 to D30, n (%) | 30 (57.7) | 25 (83.3) | 5 (35.7) | N/A |
| D31 to D100, n (%) | 14 (26.9) | 5 (16.7) | 9 (64.3) | N/A |
| Median days from first positive to first negative qPCR**, [IQR] | 11 [7-17] | 12 [7-15.5] | 11 [6-18] | 11 [7-18] |
| ***T. gondii* infection, n (%)** | 45 (86.5) | 28 (93.3) | 12 (85.7) | 5 (62.5) |
| ***T. gondii* disease, n (%)** | 7 (13.5) | 2 (6.7) | 2 (14.3) | 3 (37.5) |
| **Toxoplasmosis-attributable death, n (%)** | 0 | N/A | N/A | N/A |
| **Death-contributing TI or TD, n (%)** | 3 (5.8) | 2 (6.7) | 0 | 1 (12.5) |

Abbreviations; alloHSCT: allogeneic hematopoietic stem cell transplantation; D: donor; IQR:
interquartile range; N/A: not applicable; qPCR: quantitative polymerase chain reaction; R: recipient; TI:
*Toxoplasma* infection; TD: *Toxoplasma* disease; w/o: without; yrs: years.
\* Aplastic anemia (n= 3), primary immune deficiency (n= 1)
\*\* Three missing data

Forty-four out of 52 patients (84.6%) were qPCR-positive before D100, 30/44 (68.2%) without anti-*Toxoplasma* prophylaxis (28 TI, 2 TD) and 14/44 (31.8%) with prophylaxis (12 TI, 2 TD). The first qPCR-positive result was observed sooner after alloHSCT in patients without prophylaxis than with prophylaxis (median: 19 days [IQR: 14-25] vs. 48 days [IQR: 28-61], p= 0.0003). The median number of positive qPCR and the median time from first-positive to first-negative qPCR were not significantly different between patients with or without prophylaxis.

Eight out of 52 patients (15.4%) were qPCR-positive after D100, while receiving steroids for GVHD (n= 7) or chemotherapy for post-transplant acute lymphoblastic leukemia relapse (n= 1). In six cases out of eight (75%), qPCR was performed as part of a systematic screening for outpatients (n= 3) or inpatients (hospitalized for GVHD in two cases and sepsis in one case). In the other two out of eight cases (25%), qPCR was performed for neurological symptoms. In total after D100, five patients (63%) had TI and three (37%) had TD.

### Analysis of qPCR results according to treatment

#### Diagnosis before D100 (**Figure 1**)

Out of the 44 patients diagnosed before D100, 30 patients had no prophylaxis. Two of them (7%) became spontaneously qPCR-negative on a second evaluation, but were nevertheless subsequently started on prophylactic treatment and qPCR remained negative. For the other 28 patients, a second qPCR showed an increased parasitic load. Fifteen out of 28 patients (53.6%) received pre-emptive treatment at prophylactic dosage: all patients eventually became PCR-negative, including five after a change to non-prophylactic dosage treatment. Treatment was well tolerated without toxicity-related withdrawal. Thirteen out of 28 patients (46.4%) received pre-emptive treatment at non-prophylactic dosage: two patients with TI died of GVHD with persistently positive *Toxoplasma* qPCR, while 11 patients (nine TI and two CNS TD) became qPCR-negative (including one after change from intermediate to high dosage treatment). Treatment was changed for toxicity in three cases and suspicion of P/Sa malabsorption in one case. The detailed prescriptions and outcomes are reported in **Figure 1**.

**Figure 1:**
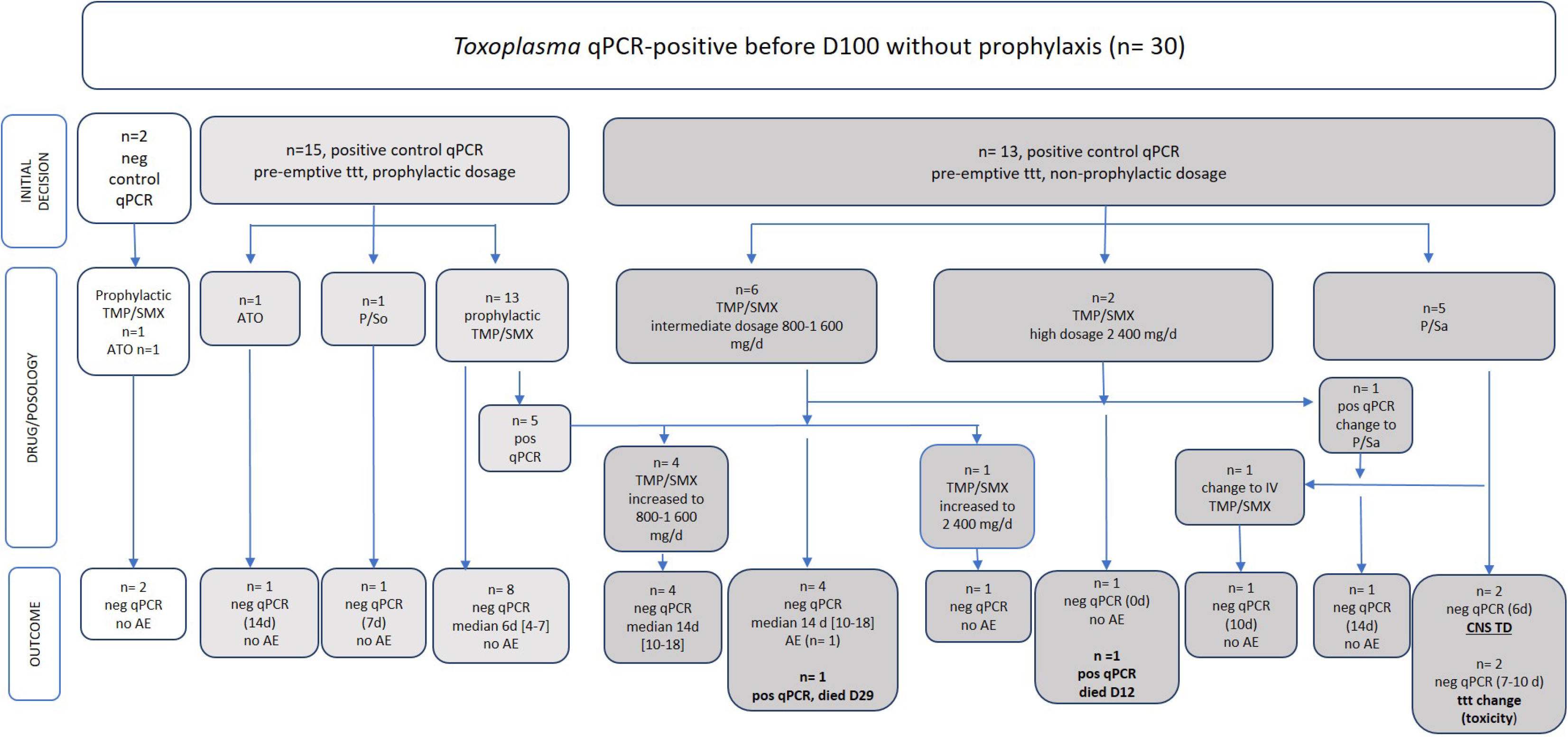
Positive *Toxoplasma* qPCR before D100 in patients without anti-*Toxoplasma* prophylaxis. Abbreviations: AE: adverse event; ATO: atovaquone; CNS: central nervous system; CR: complete remission: d: day; IV: intra-venous; neg: negative; P: pyrimethamine; pos: positive; Sa: sulfadiazine; So: sulfadoxine; TD: *Toxoplasma* disease; TMP/SMX: trimethoprim/sulfamethoxazole; ttt: treatment. Treatment length before PCR-negativity is indicated in brackets.

For the 14 qPCR-positive patients while receiving anti-*Toxoplasma* prophylaxis (**Figure 2**), the first qPCR was positive a median of 18 days [IQR: 8-34] after prophylaxis started. Prophylaxis was not modified in 8/14 (57%) TI patients, and all became PCR-negative. Prophylaxis was changed to pre-emptive non-prophylactic treatment in 6/14 (43%) cases, including two CNS TD. All patients eventually became qPCR-negative and both cases with CNS TD resolved (including one after a change of high-dosage treatment). Treatment was changed for toxicity in one case.

**Figure 2:**
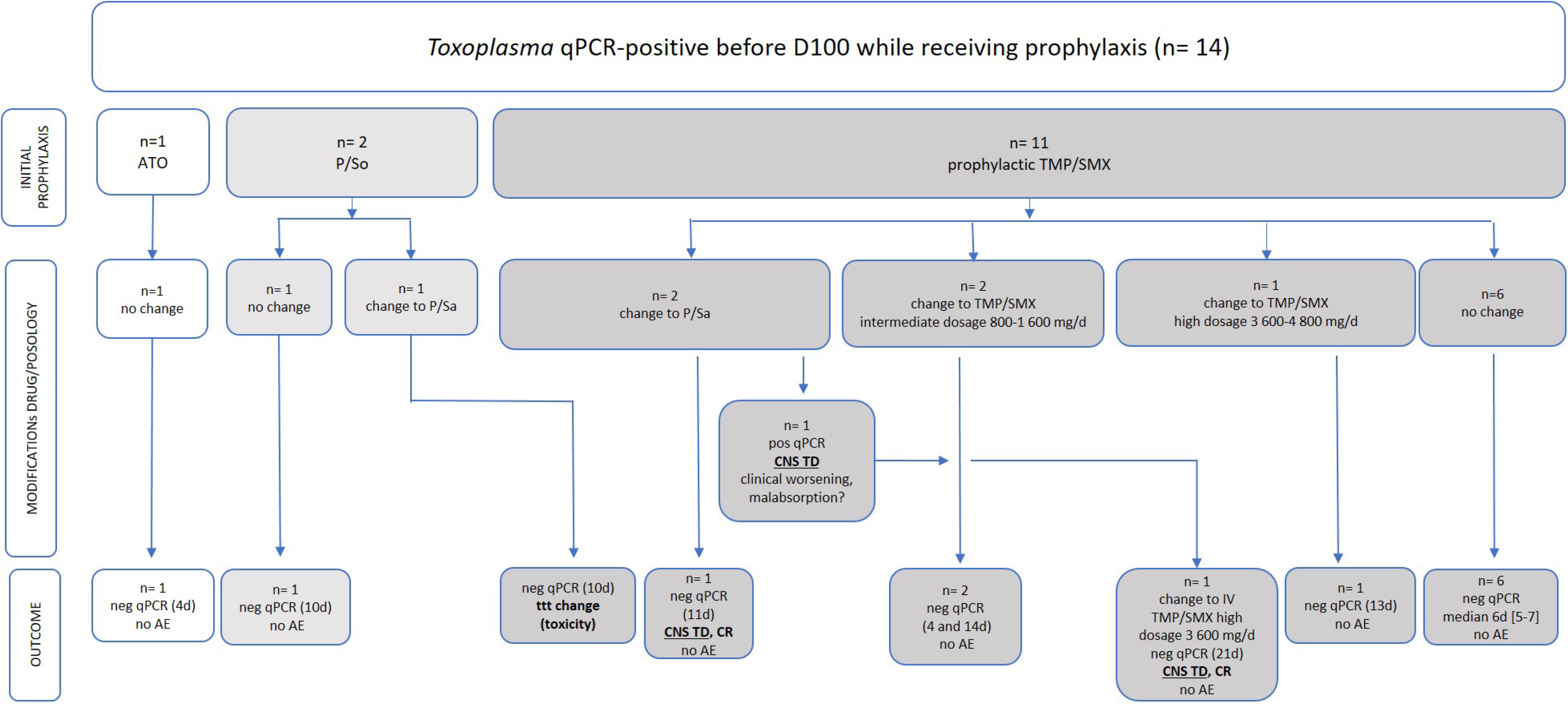
Positive *Toxoplasma* qPCR before D100 in patients receiving anti-*Toxoplasma* prophylaxis. Abbreviations: AE: adverse event; ATO: atovaquone; CNS: central nervous system; CR: complete remission: d: day; IV: intra-venous; neg: negative; P: pyrimethamine; pos: positive; Sa: sulfadiazine; So: sulfadoxine; TD: *Toxoplasma* disease; TMP/SMX: trimethoprim/sulfamethoxazole; ttt: treatment. Treatment length before PCR-negativity is indicated in brackets.

In total, all 25/44 patients (56.8%) who started or continued prophylactic dosage treatment became qPCR-negative: 20/25 (80%) received only prophylactic dosage treatment, and 5/25 (20%) needed treatment increase to non-prophylactic dosage. In total, 24/44 patients (54.5%) received non-prophylactic dosage treatment: qPCR became negative in 22/24 cases (91.7%), while TI contributed to death in two cases. No death was attributable to TI or TD.

#### Diagnosis after D100 (n=8)

*Toxoplasma gondii* qPCR was positive after D100 in eight patients (**Figure 3**). One patient with TI and one with TD without prophylaxis received pyrimethamine/sulfadiazine (P/Sa) at non-prophylactic dosage. Both patients died, and CNS TD contributed to death in one case. Breakthrough TI (n=4) or TD (n=2) occurred in six patients while receiving P/So (n= 1) or prophylactic TMP/SMX (n= 5). Three patients remained on prophylaxis and all became qPCR-negative. Three patients were changed from prophylaxis to pre-emptive treatment at non-prophylactic dosage, and all became qPCR-negative. Treatment was changed for toxicity in four cases.

**Figure 3:**
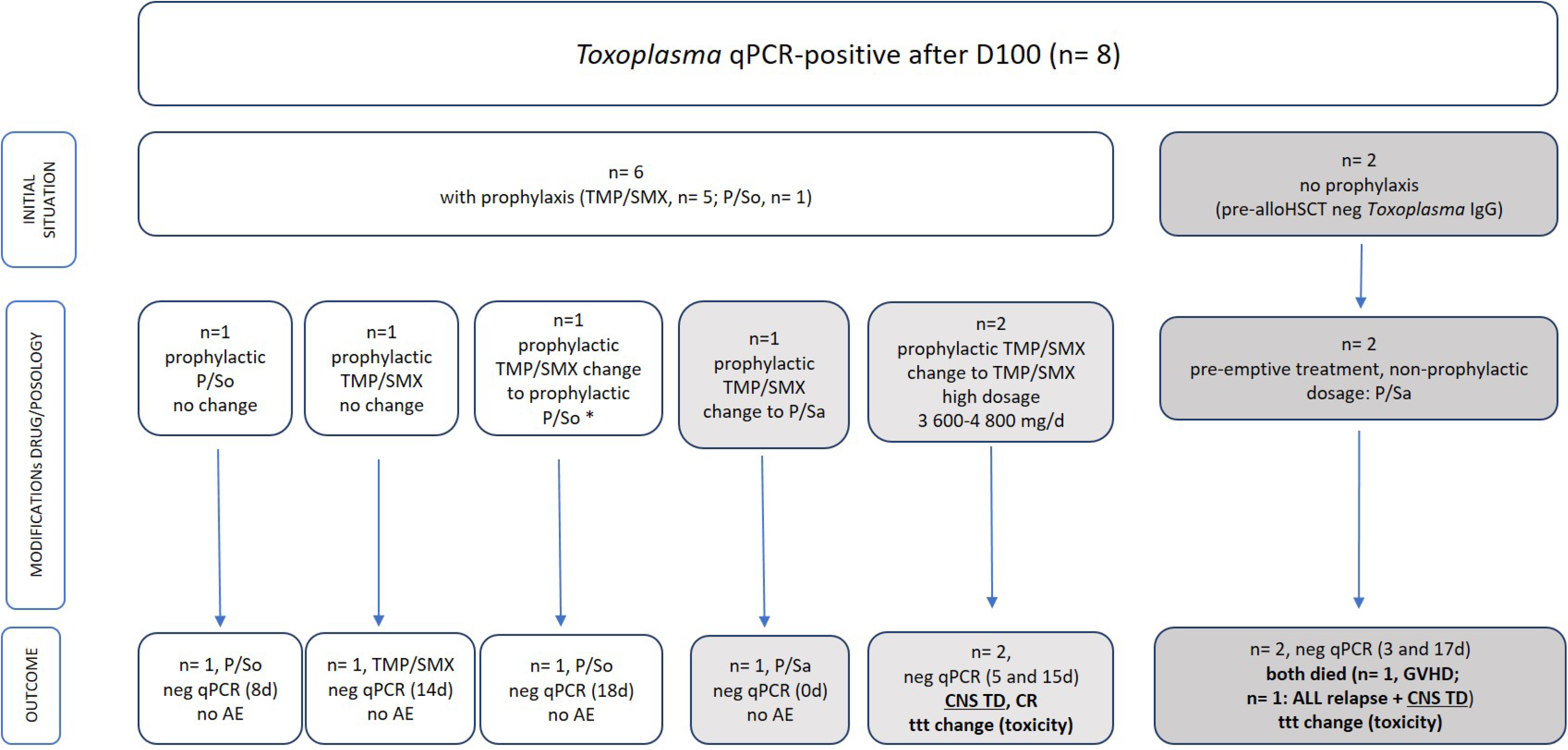
Positive *Toxoplasma* qPCR after D100. Abbreviations: ALL: acute lymphoblastic leukemia; CR: complete remission: CNS: central nervous system; d: day; GVHD: graft-versus-host-disease; P: pyrimethamine; Sa: sulfadiazine; So: sulfadoxine; TMP/SMX: trimethoprim/sulfamethoxazole; Treatment length before PCR-negativity is indicated in brackets. *TMP/SMX changed to P/So in one patient as P/So commercialization stopped at that time.

#### DNA load kinetic (**Figure 4**)

We were able to analyze the parasitic load of 24 patients with a qPCR result available either on the day of treatment implementation or the previous day, and a qPCR result within seven days before and after treatment began. Before treatment, the median decrease of the Cq values was 1 Cq/day [0.5-1.5] (i.e., a median increase of 4.0 [1.1-30.3] parasites/ml of blood/day). After treatment, assuming that the regression was linear between two points, the median increase was 1.2 Cq/day [0.4-2.3] (i.e., a median decrease of 2.8 [1.1-13.8] parasites/ml of blood/day (after calculation performed as in (14)). The results are presented in **Figure 4** according to the anti-*Toxoplasma* treatment and dosage, without a significant difference on the kinetics.

**Figure 4:**
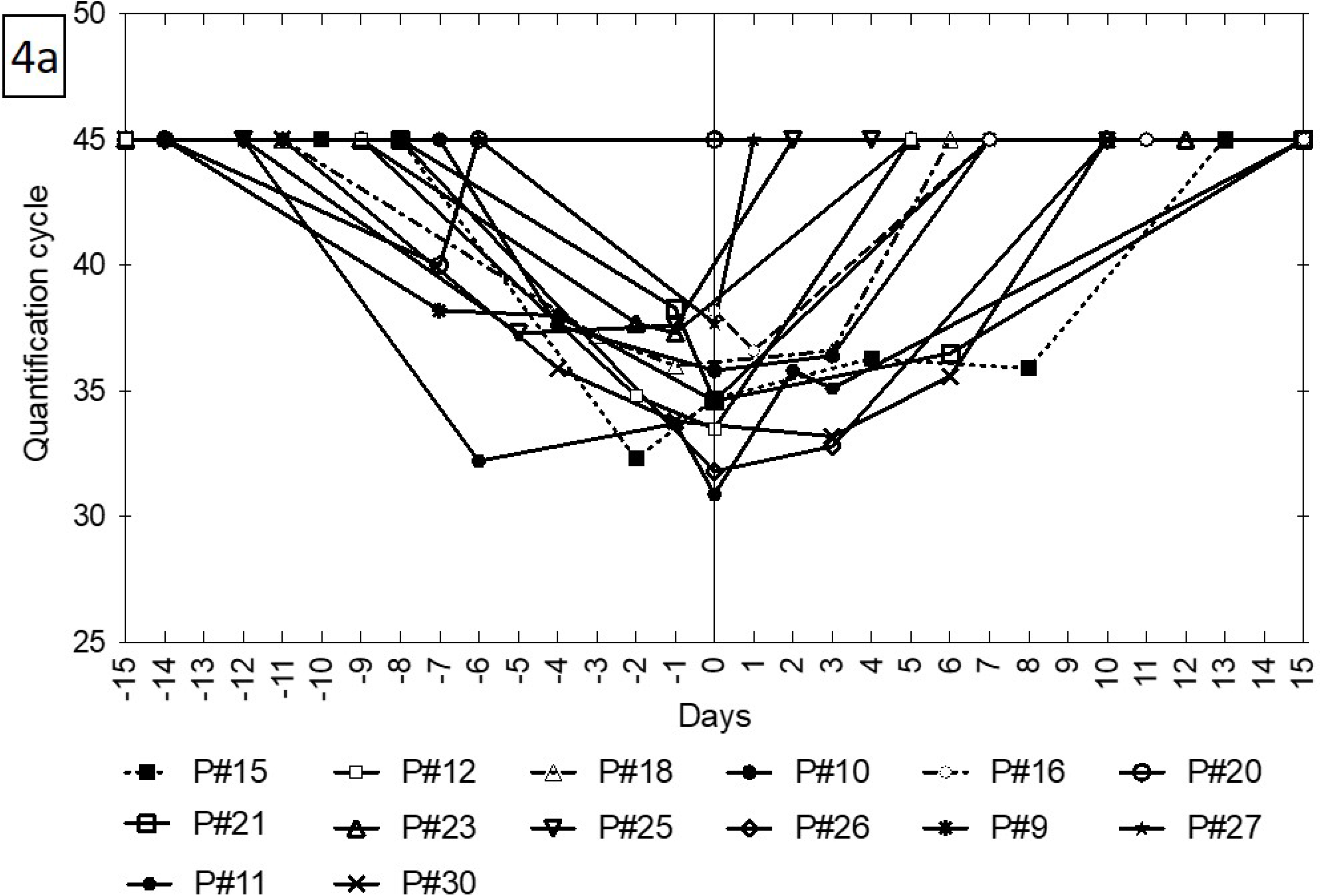
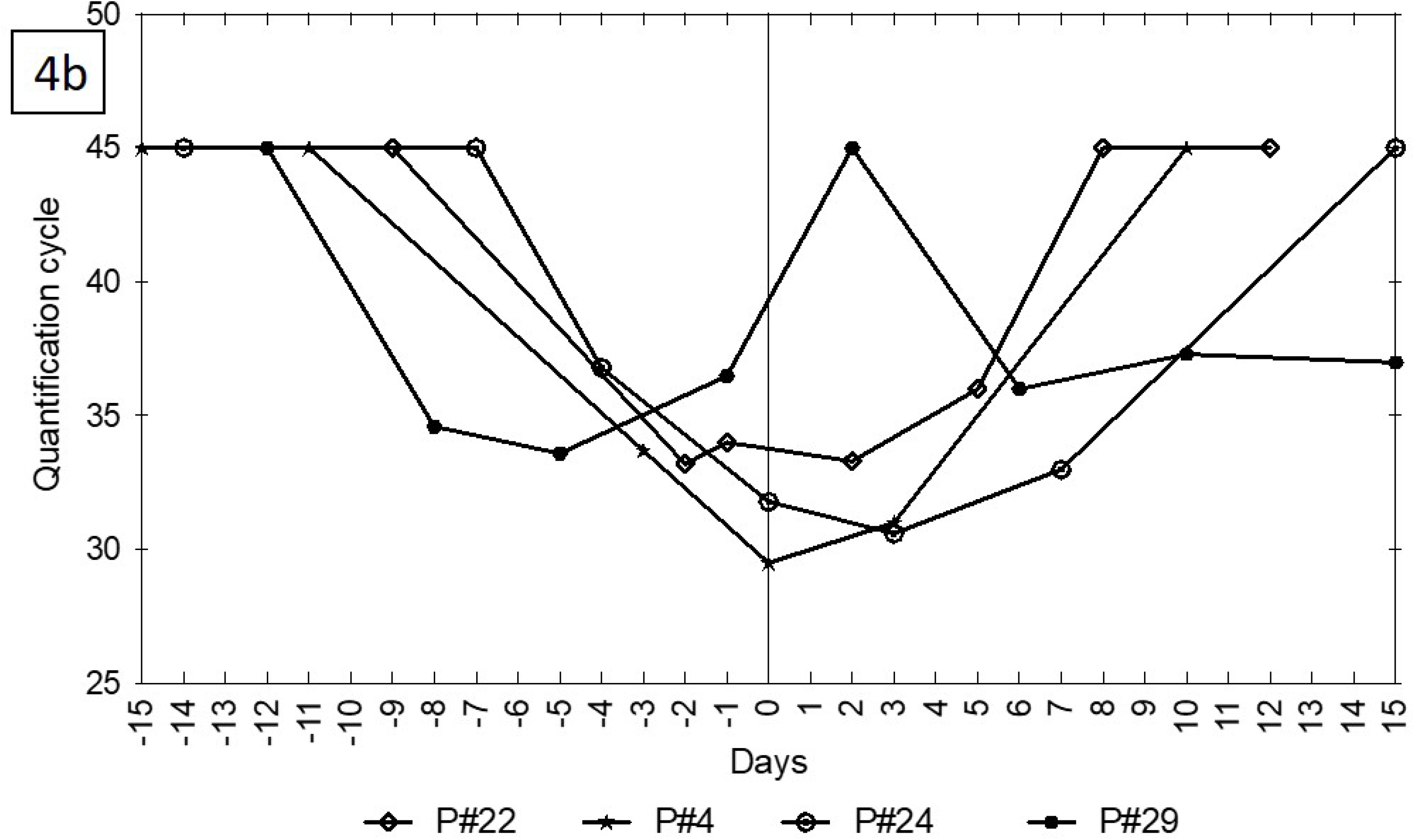
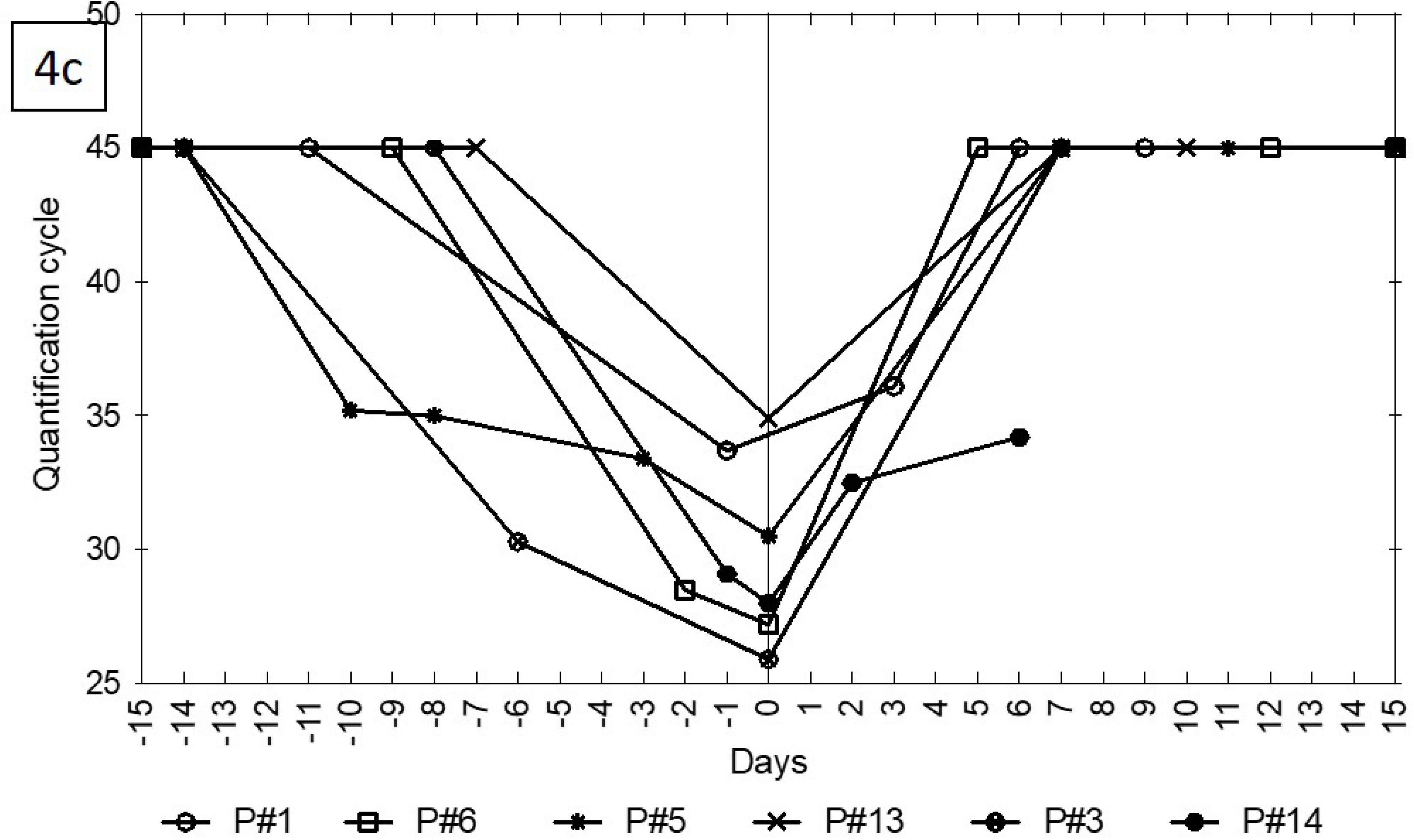
Evolution of *Toxoplasma gondii* DNA load by qPCR on blood sample in 24 patients without anti-*Toxoplasma* prophylaxis, diagnosed before D100 with TI (n= 22) or TD (n= 2). **Figure 4a: 14 patients received pre-emptive treatment at prophylactic dosage (prophylactic TMP/SMX, n= 13 and atovaquone, n= 1 (P#11))** **Figure 4b: Four patients received pre-emptive treatment at non-prophylactic dosage with intermediate dosage TMP/SMX (800-1600mg/day). P#4 died of GVHD with persistently positive qPCR** **Figure 4c: Six patients received pre-emptive treatment at non-prophylactic dosage with high dosage TMP/SMX (n= 2, P#13 and 14) or P/Sa (n= 4). P#14 died of GVHD with persistently positive qPCR.** From a starting sample with 45 Cq value corresponding to no detectable circulating DNA, patients had a decrease of Cq values (i.e. an increase of blood parasite load) until the day of pre-emptive treatment (D0), followed by an increase of Cq values (i.e. the decrease of blood parasite load) until negation or a persistent parasite load in two patients (P#29 and P#14)

## Discussion

Despite the high prevalence of anti-*Toxoplasma* IgG positivity (58.3% of alloHSCT recipients), toxoplasmosis was infrequent in our 10 year-long study, with 0.56% and 3.5% of our patients diagnosed with TI and TD, respectively. The overall prevalence of 4.13% is close to the prevalence reported in the most recent studies (3.9-4.1%) (7,18,19), whereas older studies reported higher figures, from 8.7 to 18.6% (4,6,20–22). This decrease can be ascribed to the decreasing seroprevalence in the general population, due to changes in lifestyle and food consumption (23). Other potential sources of differences include the seropositivity prevalence in the considered country (24), the screened population (only seropositive or all alloHSCT recipients (19)), the PCR assay used (the AF146527 DNA sequence being more sensitive than the B1 gene (25)), the number of positive PCR results considered for diagnosis (≥2 in (18), versus ≥1 in others), and the frequency of sampling (usually once weekly). Finally, it can also be simply ascribed to a better acceptance of TMP/SMX as prophylaxis of both toxoplasmosis and pneumocystosis (21).

In our cohort, the first positive qPCR during systematic screening was mainly observed (30/44= 68.2%) before prophylaxis started, as previously reported (7,19,21). A second qPCR with an increased parasitic load definitively advocated for an ongoing multiplication of the parasite, and therefore prompt pre-emptive treatment. Moreover, the decrease of the parasitic load following treatment implementation was indicative of drug effectiveness (26). Therefore, the qPCR screening is of utmost importance before prophylaxis is begun, or when the center policy is to avoid TMP/SMX (18).

We confirm that systematic qPCR screening is associated with a better prognosis of toxoplasmosis (6), as no death was directly attributable to toxoplasmosis in our cohort. However, the prognosis of our patients was still dismal, with an overall mortality rate of 43% in patients diagnosed before D100. This conclusion differs from that of Conrad et al., who reported a toxoplasmosis-attributable mortality of 43.5% (19). In our cohort, the decrease of the parasitic load definitively shows that the anti-*Toxoplasma* treatment was effective in controlling the parasite growth, acknowledging this is not sufficient to avoid death (GVHD was the main cause of death in our cohort, as already described (27)).

We report the occurrence of breakthrough TI and TD in 20 patients receiving prophylaxis (including TMP/SMX in 11 and five cases before and after D100, respectively), although it is hard to evaluate observance for cases occurring late after alloHSCT. Hypotheses are that the usual prophylaxis dosage is suboptimal to control the parasite, or that gut GVHD alters prophylactic treatment absorption. One can also wonder if the qPCR-positivity in these cases can be considered as a marker of global frailty and immunosuppression, placing these patients into a population at high risk of death.

If one can reach a consensus on the use of qPCR for screening alloHSCT recipients, there are no definite recommendations concerning drugs and dosage. Firstly, some patients can become spontaneously qPCR-negative. This can be expected in cases of a parasitic load at the limit of detection of the qPCR and one can wait for a second positive result before starting anti-*Toxoplasma* treatment, as proposed by Aerts et al. (18). In the case of a negative control qPCR, the hypothesis would be that these patients have cleared the infection, thanks to anti-*Toxoplasma* immune recovery. For the two patients in our case, we still decided to introduce prophylactic treatment, whereas Aerts et al. do not propose specific treatment (18). Secondly, some patients can control TI with a prophylactic dosage treatment (80% of patients in this study receiving prophylactic dosage treatment became qPCR-negative without a need for treatment increase to non-prophylactic dosage), whereas others need non-prophylactic dosage treatment. The introduction of a pre-emptive treatment at a prophylactic or intermediate dosage can be regarded as a reasonable option when associated with close clinical and qPCR monitoring integrating parasitic load quantification.

Most cases were diagnosed among donor negative/recipient positive alloHSCT, as already reported (10,13,19,21,28,29), confirming that reactivation is the main explanation for *T. gondii* infection: this raises the issue of whether or not to include the seronegative patients into the qPCR screening strategy (10,13,19,21,28–30). In our cohort, four TI and one TD occurred in pre-alloHSCT seronegative patients, i.e. a prevalence among seronegative recipients of 1%. One hypothesis for this finding is false-negative serology, due to an impaired or waning immunity, knowing that transmission of infection via the graft is also possible, although never demonstrated (10,19,28,29,31). When feasible, a negative pre-alloHSCT serology should be controlled on a previous sample, dating from the hematological malignancy diagnosis. To include or not seronegative patients in the screening is a cost-effectiveness issue that should be balanced with local seroprevalence, available financial resources and the risk of missing *Toxoplasma* disease, knowing that the seronegative population is growing (23).

The strength of our study is the large size of our cohort and the systematic screening of all alloHSCT recipients over a 10-year period, using similar serological and molecular tools. However, the retrospective nature of our study is a limit: sampling was not performed similarly in all patients, making it difficult to interpret the kinetics of the parasitic load. Similarly, therapeutic attitudes were dependent on physician decisions, in the absence of clear recommendations on the molecules and doses to be used. Despite these limits, in cases of asymptomatic TI diagnosed before the start of prophylaxis, the introduction of a pre-emptive treatment at a prophylactic or intermediate dosage could lead to *T. gondii* control: this strategy could decrease the potential toxicity of P/Sa or high-dose TMP/SMX treatments. When occurring in patients already receiving anti-*Toxoplasma* prophylaxis, one can also propose continuing prophylaxis, without necessarily increasing treatment dosage or changing molecules, with close clinical and qPCR monitoring.

In conclusion, in this large cohort of alloHSCT recipients with systematic *Toxoplasma gondii* qPCR screening until D100, we report a 4.13% prevalence of toxoplasmosis, with a no attributable mortality. It is important to start qPCR screening before engraftment, since most of our cases occurred before D30 and to continue until D100, then according to clinical suspicion. Optimal treatment dosage in asymptomatic qPCR-positive patients remains to be established, both in cases of the patient receiving prophylaxis or not. In any case, we advocate for a close monitoring of the parasitic load using qPCR to adjust therapeutic attitudes.

## Data Availability

All data produced in the present work are contained in the manuscript

## Acknowledgments

The authors wish to thank all the technical staff of the laboratory for performing the biological tests, and nurses who took take care of the patients, as well as Ms. Sylvie Lengay for data extraction and Mr. David Williams for manuscript editing.

## Author contributions

AX, AV, GS and SB designed the study protocol, conducted the search, extracting and analyzing data, interpreted results, and wrote the manuscript. DM conducted the statistical analyses. SH, JM and SB conducted and interpreted the qPCR tests. AX, AV, DM, MR, FSF, ND and RPL took care of patients. All authors read and approved the manuscript before submission.

---

**Supplementary Table 1.**
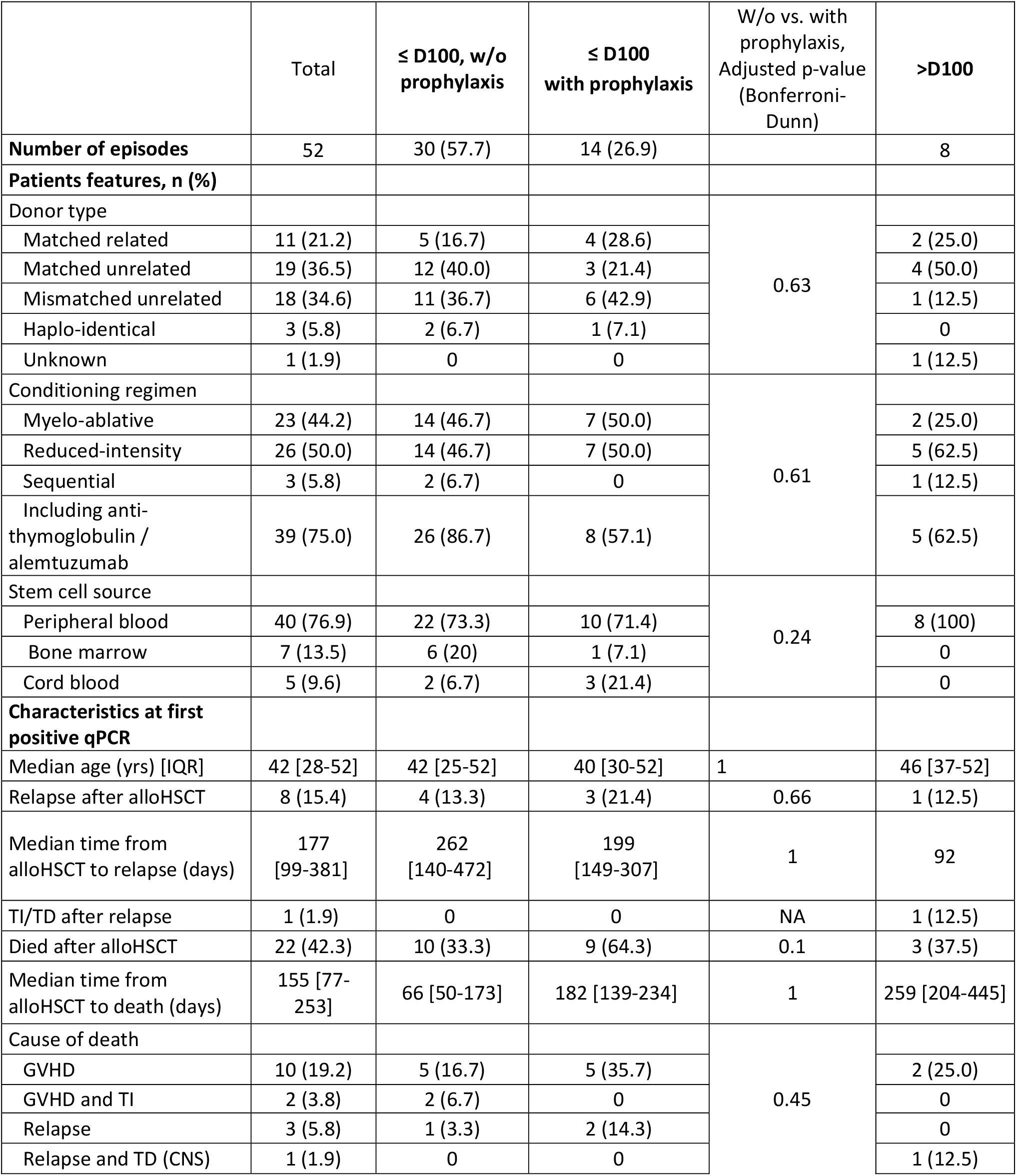
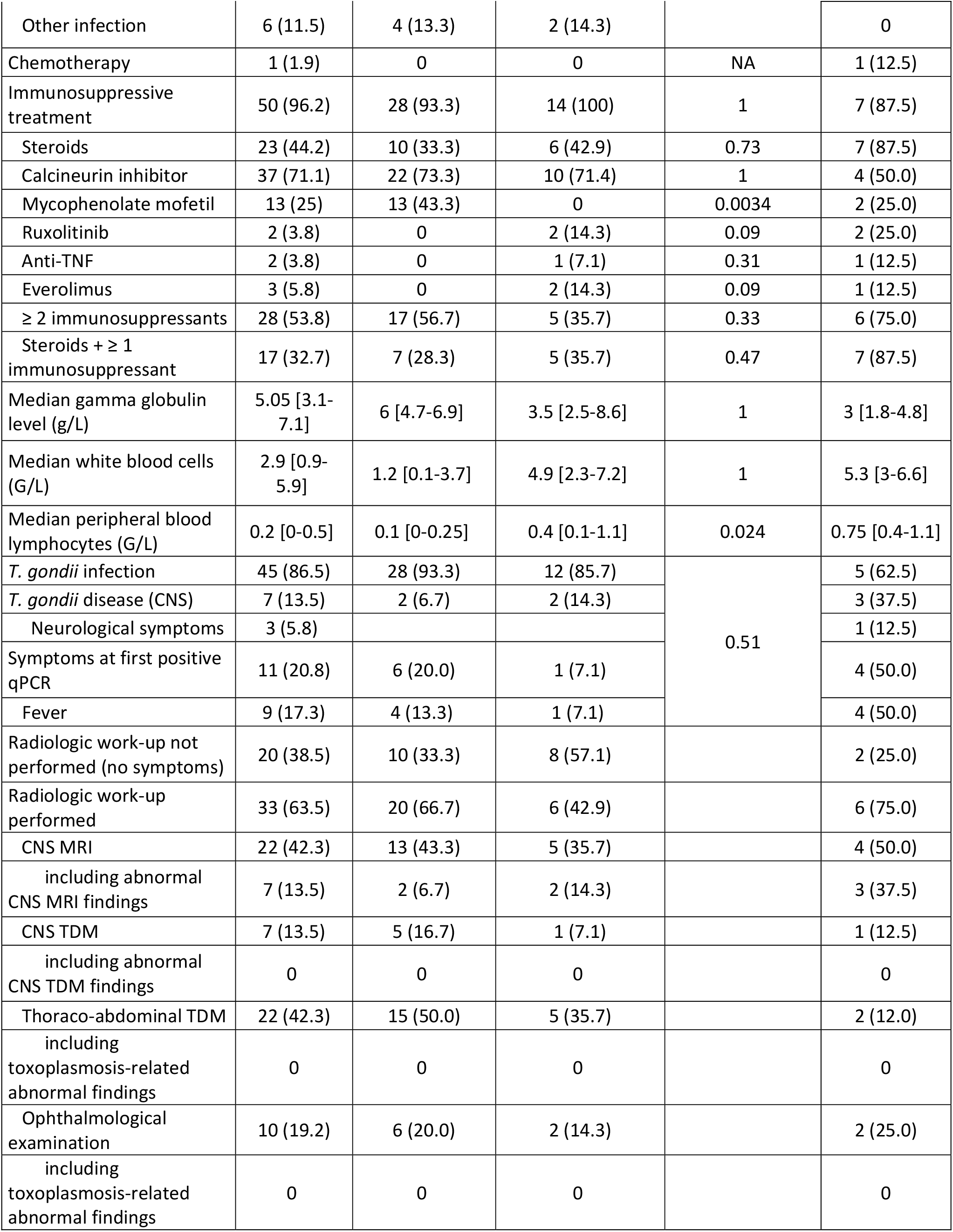
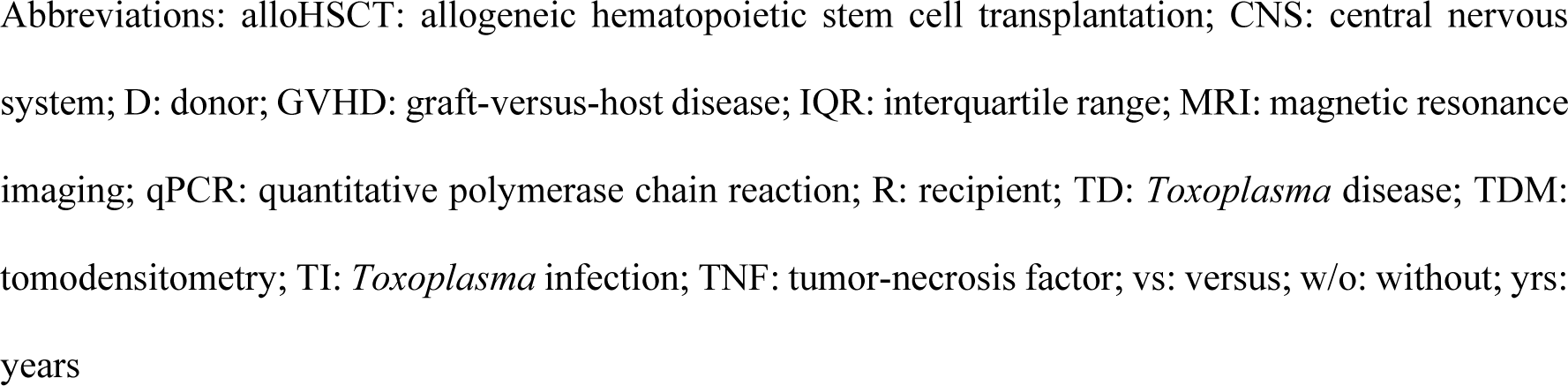
Additional characteristics of the 52 *T. gondii* positive qPCR patients according to the time of qPCR-positivity and the presence or not of prophylaxis at the first qPCR-positive result.

## References

1. Rauwolf KK, Floeth M, Kerl K, Schaumburg F, Groll AH. Toxoplasmosis after allogeneic haematopoietic cell transplantation—disease burden and approaches to diagnosis, prevention and management in adults and children. Clin Microbiol Infect. 2021 Mar;27(3):378–88.

2. Tomblyn M, Chiller T, Einsele H, Gress R, Sepkowitz K, Storek J, et al. Guidelines for Preventing Infectious Complications among Hematopoietic Cell Transplantation Recipients: A Global Perspective. Biol Blood Marrow Transplant. 2009 Oct;15(10):1143–238.

3. on behalf of the Infectious Diseases Working Party of the German Society for Hematology and Medical Oncology (AGIHO/DGHO) and the DAG-KBT (German Working Group for Blood and Marrow Transplantation), Ullmann AJ, Schmidt-Hieber M, Bertz H, Heinz WJ, Kiehl M, et al. Infectious diseases in allogeneic haematopoietic stem cell transplantation: prevention and prophylaxis strategy guidelines 2016. Ann Hematol. 2016 Sep;95(9):1435–55.

4. Martino R, Bretagne S, Einsele H, Maertens J, Ullmann AJ, Parody R, et al. Early Detection of Toxoplasma Infection by Molecular Monitoring of Toxoplasma gondii in Peripheral Blood Samples after Allogeneic Stem Cell Transplantation. Clin Infect Dis. 2005 Jan 1;40(1):67–78.

5. Lim Z, Baker B, Zuckermann M, Wade JJ, Ceesay M, Ho AYL, et al. Toxoplasmosis following alemtuzumab based allogeneic haematopoietic stem cell transplantation. J Infect. 2007 Feb;54(2):e83–6.

6. Bretagne S, Costa JM, Foulet F, Jabot-Lestang L, Baud-Camus F, Cordonnier C. Prospective study of toxoplasma reactivation by polymerase chain reaction in allogeneic stem-cell transplant recipients: T. gondii PCR for SCT recipients. Transpl Infect Dis. 2000 Sep;2(3):127–32.

7. Robin C, Leclerc M, Angebault C, Redjoul R, Beckerich F, Cabanne L, et al. Toxoplasmosis as an Early Complication of Allogeneic Hematopoietic Cell Transplantation. Biol Blood Marrow Transplant. 2019 Dec;25(12):2510–3.

8. De Medeiros BC, De Medeiros CR, Werner B, Loddo G, Pasquini R, Bleggi-Torres LF. Disseminated toxoplasmosis after bone marrow transplantation: report of 9 cases: Disseminated toxoplasmosis after BMT. Transpl Infect Dis. 2001 Mar;3(1):24–8.

9. Mele A, Paterson P, Prentice H, Leoni P, Kibbler C. Toxoplasmosis in bone marrow transplantation: a report of two cases and systematic review of the literature. Bone Marrow Transplant. 2002 Apr 1;29(8):691–8.

10. Derouin F, Devergie A, Auber P, Gluckman E, Beauvais B, Garin YJF, et al. Toxoplasmosis in Bone Marrow-Transplant Recipients: Report of Seven Cases and Review. Clin Infect Dis. 1992 Aug 1;15(2):267–70.

11. Matsuo Y, Takeishi S, Miyamoto T, Nonami A, Kikushige Y, Kunisaki Y, et al. Toxoplasmosis encephalitis following severe graft-vs.-host disease after allogeneic hematopoietic stem cell transplantation: 17 yr experience in Fukuoka BMT group: Toxoplasmosis encephalitis after allo-HSCT. Eur J Haematol. 2007 Oct;79(4):317–21.

12. Redjoul R, Robin C, Foulet F, Leclerc M, Beckerich F, Cabanne L, et al. Pneumocystis jirovecii pneumonia prophylaxis in allogeneic hematopoietic cell transplant recipients: can we always follow the guidelines? Bone Marrow Transplant. 2019 Jul;54(7):1082–8.

13. Martino R, Maertens J, Bretagne S, Rovira M, Deconinck E, Ullmann AJ, et al. Toxoplasmosis after Hematopoietic Stem Cell Transplantation. Clin Infect Dis. 2000 Nov 15;31(5):1188–94.

14. Menotti J, Garin YJF, Thulliez P, Sérugue MC, Stanislawiak J, Ribaud P, et al. Evaluation of a new 5’-nuclease real-time PCR assay targeting the Toxoplasma gondii AF146527 genomic repeat. Clin Microbiol Infect. 2010 Apr;16(4):363–8.

15. Carreras E, Dufour C, Mohty M, Kröger N, editors. The EBMT Handbook: Hematopoietic Stem Cell Transplantation and Cellular Therapies [Internet]. Cham: Springer International Publishing; 2019 [cited 2022 Sep 26]. Available from: http://link.springer.com/10.1007/978-3-030-02278-5

16. Bustin SA, Benes V, Garson JA, Hellemans J, Huggett J, Kubista M, et al. The MIQE Guidelines: Minimum Information for Publication of Quantitative Real-Time PCR Experiments. Clin Chem. 2009 Apr 1;55(4):611–22.

17. Foot AB, Garin YJ, Ribaud P, Devergie A, Derouin F, Gluckman E. Prophylaxis of toxoplasmosis infection with pyrimethamine/sulfadoxine (Fansidar) in bone marrow transplant recipients. Bone Marrow Transplant. 1994 Aug;14(2):241–5.

18. Aerts R, Mercier T, Beckers M, Schoemans H, Lagrou K, Maertens J. Toxoplasmosis after allogeneic haematopoietic cell transplantation: experience using a PCR-guided pre-emptive approach. Clin Microbiol Infect. 2022 Mar;28(3):440–5.

19. Conrad A, Le Maréchal M, Dupont D, Ducastelle-Leprêtre S, Balsat M, Labussière-Wallet H, et al. A matched case–control study of toxoplasmosis after allogeneic haematopoietic stem cell transplantation: still a devastating complication. Clin Microbiol Infect. 2016 Jul;22(7):636–41.

20. Fricker-Hidalgo H, Bulabois C, Brenier-Pinchart M, Hamidfar R, Garban F, Brion J, et al. Diagnosis of Toxoplasmosis after Allogeneic Stem Cell Transplantation: Results of DNA Detection and Serological Techniques. Clin Infect Dis. 2009 Jan 15;48(2):e9–15.

21. Paccoud O, Guitard J, Labopin M, Surgers L, Malard F, Battipaglia G, et al. Features of Toxoplasma gondii reactivation after allogeneic hematopoietic stem-cell transplantation in a high seroprevalence setting. Bone Marrow Transplant. 2020 Jan;55(1):93–9.

22. Meers S, Lagrou K, Theunissen K, Dierickx D, Delforge M, Devos T, et al. Myeloablative Conditioning Predisposes Patients for Toxoplasma gondii Reactivation after Allogeneic Stem Cell Transplantation. Clin Infect Dis. 2010 Apr 15;50(8):1127–34.

23. Guigue N, Léon L, Hamane S, Gits-Muselli M, Le Strat Y, Alanio A, et al. Corrigendum: Continuous Decline of Toxoplasma gondii Seroprevalence in Hospital: A 1997–2014 Longitudinal Study in Paris, France. Front Microbiol. 2018 Nov 27;9:2814.

24. Foroutan-Rad M, Majidiani H, Dalvand S, Daryani A, Kooti W, Saki J, et al. Toxoplasmosis in Blood Donors: A Systematic Review and Meta-Analysis. Transfus Med Rev. 2016 Jul;30(3):116– 22.

25. Reischl U, Bretagne S, Krüger D, Ernault P, Costa JM. Comparison of two DNA targets for the diagnosis of Toxoplasmosis by real-time PCR using fluorescence resonance energy transfer hybridization probes. BMC Infect Dis. 2003 Dec;3(1):7.

26. Isa F, Saito K, Huang YT, Schuetz A, Babady NE, Salvatore S, et al. Implementation of Molecular Surveillance After a Cluster of Fatal Toxoplasmosis at 2 Neighboring Transplant Centers. Clin Infect Dis. 2016 Aug 15;63(4):565–8.

27. Bretagne S, Costa JM, Kuentz M, Simon D, Vidaud M, Fortel I, et al. Late toxoplasmosis evidenced by PCR in a marrow transplant recipient. Bone Marrow Transplant. 1995 May;15(5):809–11.

28. Chandrasekar P, Momin F, The Bone Marrow Transplant Team. Disseminated toxoplasmosis in marrow recipients: a report of three cases and a review of the literature. Bone Marrow Transplant. 1997 Apr 1;19(7):685–9.

29. Gajurel K, Dhakal R, Montoya JG. Toxoplasma prophylaxis in haematopoietic cell transplant recipients: a review of the literature and recommendations. Curr Opin Infect Dis. 2015 Aug;28(4):283–92.

30. Štajner T, Vujić D, Srbljanović J, Bauman N, Zečević Ž, Simić M, et al. Risk of reactivated toxoplasmosis in haematopoietic stem cell transplant recipients: a prospective cohort study in a setting withholding prophylaxis. Clin Microbiol Infect. 2022 May;28(5):733.e1-733.e5.

31. Dupont D, Fricker-Hidalgo H, Brenier-Pinchart MP, Garnaud C, Wallon M, Pelloux H. Serology for Toxoplasma in Immunocompromised Patients: Still Useful? Trends Parasitol. 2021 Mar;37(3):205–13.

